# Understanding the political economy of reforming global health initiatives – insights from global and country levels

**DOI:** 10.1101/2024.10.04.24314895

**Authors:** Sophie Witter, Natasha Palmer, Rosemary James, Shehla Zaidi, Severine Carillon, Rene English, Giulia Loffreda, Emilie Venables, Shifa Salman Habib, Jeff Tan, Fatouma Hane, Maria Paola Bertone, Seyed-Moeen Hosseinalipour, Valery Ridde, Asad Shoaib, Adama Faye, Lilian Dudley, Karen Daniels, Karl Blanchet

**Affiliations:** Queen Margaret University, Edinburgh, Scotland; Geneva Centre of Humanitarian Studies, Faculty of Medicine, University of Geneva, Switzerland; Aga Khan University International (in the UK) with Aga Khan University Pakistan; Cheikh Anta Diop University, Dakar, Senegal; Division of Health Systems and Public Health, Department of Global Health, Stellenbosch University, South Africa; Institut de Recherche pour le Développement, France

**Keywords:** Global health initiatives, political economy analysis, governance, financing, South Africa, Pakistan, Senegal

## Abstract

**Introduction:** Since 2000, the number and role of global health initiatives has been growing, with these platforms playing an increasingly important role in pooling and disbursing funds dedicated to specific global health priorities. While recognising their important contribution, there has also been a growth in concerns about distortions and inefficiencies linked to the GHIs and attempts to improve their alignment with country health systems. There is a growing momentum to adjust GHIs to the current broader range of global health threats, such as non-communicable diseases, humanitarian crises and climate change. However, these reform attempts hit up against political economy realities of the current structures.

**Methods:** In this article, we draw on research conducted as part of the Future of Global Health Initiatives process. The study adopted a cross-sectional, mixed-methods approach, drawing from a range of data sources and data collection methods, including a global and regional level analysis as well as three embedded country case studies in Pakistan, South Africa and Senegal. All data was collected from February to July 2023. 271 documents were analysed in the course of the study, along with data from 335 key informants and meeting participants in 66 countries and across a range of constituencies. For this paper, data were analysed using a political economy framework which focused on actors, context (especially governance and financing) and framing.

**Findings:** In relation to actors, the GHIs themselves have become increasingly complex (internally and in their interrelations with other global health actors and one another). They have a large range of clients (including at national level and amongst multilateral agencies) which function as collaborators as well as competitors. Historically there have been few incentives within any of the actors to maximise collaboration given the competitive funding landscape. Power to exert pressure for reforms sits ultimately with bilateral and private funders, though single-issue northern NGOs are also cited as important influencers. Funders have not collaborated to enable reforms, despite concerns amongst a number of them, because of the helpful functional role of GHIs, which serves funder interests. Some key global boards are reported to be engineered for stasis, and there are widespread concerns about lack of transparency and over-claiming (by some GHIs) of their results. Narratives about achievements and challenges are important to enable or block reforms and are vigorously contested, with stakeholders often selecting different outcomes to emphasise in justifying positions.

**Conclusion:** GHIs have played an important role in the global health ecosystem but despite formal accountability structures to include recipient governments, substantive accountability has been focused upwards to funders, with risk management strategies which prioritise tracking resources more than improved national health system performance. Achieving consensus on reforms will be challenging but funding pressures and new threats are creating a sense of urgency, which may shift positions. Political economy analysis can model and influence these debates.

## Introduction

The global health system has undergone significant expansion over the past few decades, linked in part to efforts to reach the Millennium Development Goals (MDGs). This has included a continued increase in both the number and diversity of actors and the volume of funding. It is of note that there has also been a marked increase in the distribution of development assistance for health (DAH) through Global Health Initiatives (GHIs), which are international partnerships that aim to address specific goals in global health. Many GHIs have been established since the early 2000s, driven by the creation of the Global Fund to fight AIDS, Tuberculosis and Malaria (GFATM) and Gavi (the Vaccine Alliance), which accounted for 14% of DAH by 2019 (1). Four “mega- trends” in DAH of proliferation, verticalization, circumvention of government systems, and fragmentation are identified, which go beyond but include the role of the GHIs. In relation to health financing, it is also important to note that DAH still forms a large part of the health budgets for many low-income countries (LICs) in particular, and yet that the overall amount of financing for health is not adequate to fund the achievement of the Sustainable Development Goals (SDGs). (2)

Emerging challenges such as climate change, humanitarian crises, antimicrobial resistance, and a rise in non-communicable diseases over this timescale also suggest a need to find ways of approaching global health which are less vertically focussed on infectious diseases. Plateauing DAH and shrinking fiscal space post-COVID-19, a stormy geopolitical context, growing health needs and costly health technologies are amongst the additional expected stressors. These factors argue for an urgent review to ensure that all global health resources are used as effectively as possible. The mismatch of DAH overall to global and country burden of disease suggests scope for improvement.

The FGHI process was a time-bound multi-stakeholder exercise to explore how Global Health Initiatives (GHIs) contribute to progress towards Universal Health Coverage (UHC) and the broader SDGs 2030 Agenda, and how this could be strengthened from the perspective of recipient countries. The process, which ran 2022-23, aimed to make recommendations on how GHIs could be more efficient, effective and equitable and to catalyse collective action to ensure that they were fit for purpose through 2030 and beyond. It led to the endorsement of the Lusaka Agenda in December 2023, which outlines five key shifts and a call to action for all GHI stakeholders to strengthen the contribution of GHIs to achieving UHC (3).

This article draws from research commissioned as an input into that process (4). It was focused on six GHIs, which differ in form and function: the GFATM, Gavi, the Global Financing Facility for Women, Children, and Adolescents (GFF), Unitaid, the Foundation for Innovative New Diagnostics (FIND), and the Coalition for Epidemic Preparedness Innovations (CEPI) (Table 1), however in this article we focus on the three main GHIs which account for the majority of funding invested in low- and middle-income countries (GFATM, Gavi and the GFF). The study adopted a UHC lens and focused on countries’ experiences with the GHIs as a group and the wider aid ecosystem.

**Table 1.** The six Global Health Initiatives selected for the FGHI study.

| Global health initiative (GHI) | Main objective | Country-level function | Approximate size |
| --- | --- | --- | --- |
| **Country-level grants and technical assistance** |  |  |  |
| **The Global Fund to Fight AIDS, Tuberculosis and Malaria (GFATM)**<br>Est. 2002.<br>Headquartered in Geneva | To attract leverage and invest additional resources to end epidemics of HIV, TB, malaria, reduce health inequities and support attainment of the SDGs | Grants and technical assistance for disease programmes and health system strengthening relating to these programmes. | \$5.2 billion per year<sup>1</sup>. (67)<br>Country eligibility is based on income classification and disease burden of HIV, TB, and/or |

<sup>1</sup> By taking the last replenishment total and dividing by the three-year cycle; not a measure of actual expenditure per year

|  |  |  |  |
| --- | --- | --- | --- |
|  |  |  | malaria. |
| **Gavi, the vaccine alliance**<br>Est. 2000.<br>Headquartered in Geneva. | To save lives and increase people's health by increasing the equitable and sustainable use of vaccines | Grants and technical assistance for vaccination programmes and health system strengthening relating to these programmes. | US\$21.3 billion in donor contributions and pledges from 2021-2025.(68)<br>Country eligibility depends on Gross National Income per capita. |
| **Model based on leveraging concessional finance** |  |  |  |
| **Global Financing Facility (GFF)**<br>Est. 2015.<br>Headquartered in Washington, D.C. | To end all preventable maternal, child and adolescent deaths by 2030, through a health systems strengthening approach | Grants (as seed funding) and technical assistance rooted in a broad investment case, rooted through government systems. | As of June 30, 2020, the GFF Trust Fund had US\$602 million in grants under implementation—linked to US\$4.7 billion of World Bank IDA/IBRD financing<br>Aims to mobilize more than US\$57 billion from 2015 to 2030 (69) |

| Research & development and market shaping |  |  |  |
| --- | --- | --- | --- |
| **Unitaid**<br>Est. 2006.<br>Headquartered in Geneva | 1-To accelerate the introduction and adoption of key health products<br>2-To create systemic conditions for sustainable, equitable access<br>3-To foster inclusive and demand-driven partnerships for innovation | Global (late stage) research & development (R&D) and implementation for new innovations, including creation of sustainable market conditions for equitable access. | Portfolio budget of US\$164 million in 2023.(70)<br>Requested US\$1.5 billion for the 2023-2027 investment case.(71) |
| **FIND, the global alliance for diagnostics**<br>Est. 2003.<br>Headquartered in Geneva. | To drive equitable access to reliable diagnosis through collective action | Global R&D for new diagnostics | Requested US\$100–120 million per year for 2021-2023 (72) |
| **Coalition for Epidemic Preparedness Innovations (CEPI)**<br>Est. 2016.<br>Headquartered in London. | To accelerate the development of vaccines and other biologic countermeasures against epidemic and pandemic threats to be accessible to all | Global R&D for new vaccines and other measures to prevent epidemics and pandemics | Approximately \$200 million per year.<br>Overall target of funds of USD \$1billion. (73) |

This article reports on the political economy underlying the current role of GHIs in the global health system and attempts to reform them. While critiques of GHIs have been expressed and published for decades (5–8) and incremental reforms undertaken within organisations, reforming fundamental aspects such as mandates, governance, transparency and priorities, and how GHIs and other DAH actors cooperate with one another and engage with national health systems, has been challenging.

## Materials and methods

The study adopted a cross-sectional, mixed-methods approach, drawing from a range of data sources and data collection methods, including a global and regional level analysis as well as three embedded country case studies in Pakistan, South Africa and Senegal. Case study countries were selected based on offering a range of national government’s experiences with GHIs, having a variety of GHIs’ investments and having in-country strong academic partners.

### Data sources

The study was conducted between February and July 2023 and drew on a number of data sources, which are detailed more fully in (4) : 1) a rapid scoping review of available peer-reviewed and grey literature (271 documents in total), 2) global and country level burden of disease and health financing data, 3) global-level key informants (KIs) interviews, 4) three in-depth country case studies, 5) regional consultations with key stakeholders in all six World Health Organization (WHO) regions, 6) an online survey targeted to KIs who could not join the interviews or consultations and Board members of the GHIs, and 7) consultative meetings, including one co-hosted by the Africa Centre for Disease Control and Prevention (CDC) in Addis Ababa in June 2023 to discuss preliminary findings. The study participants (total of 335) were based in 66 countries (Table 2).

**Table 2.** Number and category of study participants by data source.

| Data Stream | Number of participants | Category of participants |
| --- | --- | --- |
| **Global-level interviews** | 76 | GHI (n=18), Academic (n=11), Multilateral (n=16), Bilateral donor (n=15), CSO (n=10), Private Sector (n=4), Foundation (n=2) |
| **Country-level interviews (Pakistan, Senegal, South Africa)** | 63 | Government (n=22), CSO (n=10), Academic (n=10), Implementation partner (n=4), Technical/Financial partner (n=6), National and provincial disease programme (n=4), Technical Assistance provider (n=1), Multilateral (n=3), Regional organisation (n=2), Private Sector (n=1) |
| **Regional consultations (all six WHO regions)** | 77 | Multilateral (n=23), CSO (n=23), Implementing government (n=17), Academic (n=11), Implementation partner (n=3) |
| **Product Development Partnership Coalition Consultation** | 6 | Product development partnership member (n=6) |
| **Targeted online survey** | 46 | Academic (n=15), CSO (n=11), GHI (n=6), Implementing government (n=4), Bilateral donor (n=4), Multilateral (n=4), Foundation (n=1), Other (n=2) |
| **Hybrid Deliberative Discussion co-hosted by Africa CDC** | 45<br>(30 in-person, 15 online) | *In-person:* Government (n=9), FGHI (n=4), CSO (n=4), Multilateral, (n=3), Regional organization (n=3), Africa CDC (n=3), Bilateral donor (n=2), Foundation (n=2)<br>*Online:* CSO (n=2), Product development partnership (n=1), Government (n=2), Foundation (n=5), Bilateral donor (n=2), Independent global health consultant from the African continent (n=1), Multilateral (n=1), Academic (n=1) |
| **FGHI Steering Group Consultative Meeting** | 22 | Multilateral (n=2), Recipient government (n=3), CSO (n=2), Bilateral donor (n=8), Foundation (n=5), FGHI (n=2) |
| **Total number of study participants\*** | **335** | CSO (n=62, 19%)<br>Government (n=57, 17%)<br>Multilateral (n=52, 16%)<br>Academic (n=48, 14%)<br>Bilateral donor (n=31, 9%)<br>GHI (n=24, 7%)<br>Foundation (n=15, 4%)<br>PDP (n=7, 2%)<br>FGHI (n = 6, 2%)<br>Private Sector (n=5, 1%)<br>Other (n=29, 8%) |

\*some participants may have been counted twice (e.g. if they participated in both an interview and a consultation)

Study participants were purposely selected based on their level of experience working with GHIs and their membership of relevant constituencies (GHIs, academia, multilateral or bilateral donors, civil society organizations (CSOs), private sector and philanthropic foundations). A first list of informants was drafted by the FGHI Secretariat and then completed by the professional network of the research consortium. During the course of the study, new KIs were recruited based on suggestions from people interviewed (snowball technique).

### Data analysis

All data sources were synthesised to inform this paper. The qualitative data were recorded, transcribed, and coded inductively and deductively by a team of three researchers trained in qualitative research. The researcher consortium convened frequently to discuss the emerging findings, and during analysis examined similarities and differences among GHIs and across participant categories. Political economy analysis (PEA)(9–12) was used throughout the study to inform the analysis and synthesis. Such an approach allowed the team to reflect on the dynamic interaction between actors, their relative power and respective interests and incentives, and elements of the broader context, and how the outcome of the interaction affects the likelihood and content of future changes. In particular, the study focused on analysing actors, context and framing related to the GHIs and the wider global health ecosystem (Figure 1).

**Figure 1.**
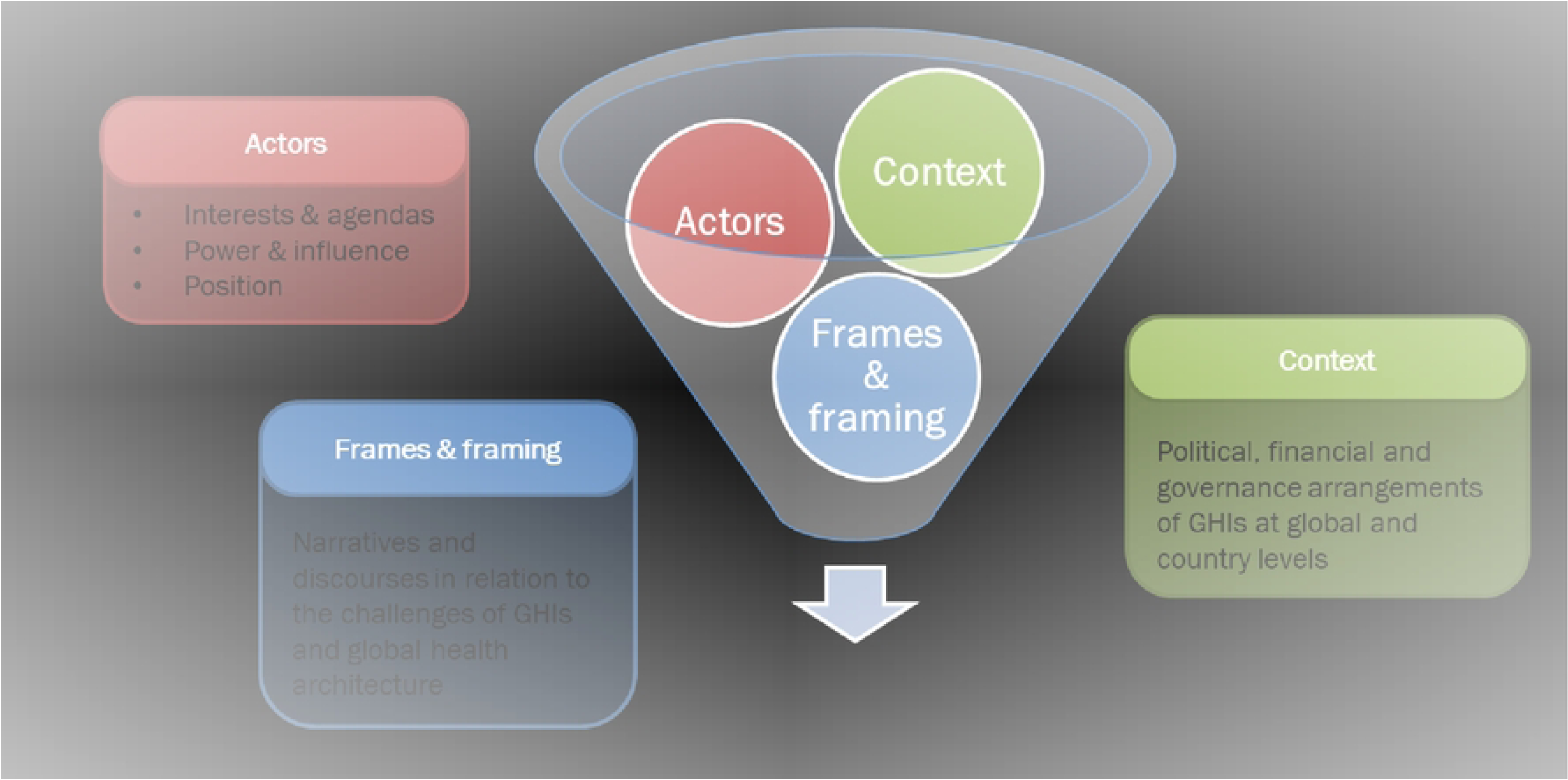
Political economy framework guiding the qualitative analysis and synthesis. *Source: Adapted from* (11)

- Actors: a detailed analysis of the stakeholders was carried out in each country and at global level. We identified as domains for the stakeholder analysis: (i) interest and position in relation to changes whether the stakeholder supports, opposes or is neutral about changes to status quo on GHIs and its motivations for this; and (ii) power and influence (i.e., the potential ability of the stakeholder to affect implementation of changes to status quo). The stakeholder analysis was informed by guidance (13–15).
- Context: we collected and analysed information concerning the broader context in which the stakeholders operate and how it can constrain or support change, focusing on governance structures and financial elements, which emerged from analysis as most relevant.
- Framing: building on recent literature (2) which acknowledges the critical influence of frames and framing in policy processes, we explored (but in less depth) the role and power of narratives and discourses, and how they shape the debate around GHIs.

### Ethical considerations

The study was approved by the ethics review boards of University of Geneva, Cheikh Anta Diop University, Stellenbosch University, and Aga Khan University, Pakistan. Informed consent (oral and written, according to the circumstances) was obtained from the study participants to participate, and to record the qualitative data, which was pseudonymised to protect the study participants from being identified.

### Study limitations

The study set out to capture the views of highly expert stakeholders with deep insights into the workings of the GHIs, but also different perspectives on the topic, representing all the key parts of the global health system. It is important to note several limitations in this work, largely as a result of a tight timeframe. The data we collected were qualitative and based on interviews, consultations and a rapid non-systematic literature review. It is also important to highlight that this is a contested area, and there were conflicting positions, which we reflect in this article.

The country case studies were not meant to be a representative sample, but rather chosen due to strong research partnerships within the country, as well as to compare a range of contexts in which the GHIs of focus are active. Findings of one country are not meant to be generalisable to other contexts, but to shed light on the dynamics that occur around GHIs and different experiences of country stakeholders.

## Results

### Actors

There has been a significant increase in the number and diversity of actors within the system (16). Whilst 30 years ago, it comprised primarily of bilateral and multilateral arrangements between nation-states, it is now a varied landscape, which also includes private firms, philanthropies, non-governmental organisations (NGOs) and GHIs (17). The increase in DAH disbursements from 1990-2015 was accompanied by a five-fold increase in the number of actors involved in global health, with a particularly rapid rate of growth in the number of CSOs between 2005-2011 (17). In addition, there has been a marked increase in the distribution of DAH through GHIs, driven by the creation of the GFATM and Gavi (1).

There have also been changes to the GHI’s funding to partners: recent analysis suggested that GFATM’s share of disbursements to governmental organisations has been declining, from 80 percent in 2003 to 40 percent of all disbursements in 2021 (18). Many of the CSOs funded are focussed in specific health areas: separate work has found that over one-third of CSO channels are only providing funds for the implementation of programmes in one health area e.g. HIV/AIDS, malaria, child and maternal health or nutrition (17).

Over recent decades, many GHIs have grown rapidly and become major players in the global health system. They are active at global, regional and country level. Some of the longest-standing GHIs such as GFATM and Gavi have evolved into large and complex organisations with the size of their secretariats reflecting this institutional growth. They have inevitably developed their own internal dynamics and priorities. GHIs now raise and channel 14% of DAH (1,19) and have taken on a growing range of roles, most recently including COVID-19 responses.

Key stakeholder groups involved in this ecosystem include:

- **GHIs,** which are instrumental in creating and responding to specific agendas by mobilising funding and collective action. Within the GHIs themselves, it is useful to distinguish several potential loci of power and influence. The Boards are the official mechanism of governance, but other parts of the organisations such as the Secretariats or technical teams can also be important actors. In the case of the GFATM, for example, there are other bodies which act independently, such as the Office of the Inspector General and the Technical Review Panel and Technical Evaluation Reference Group, which has since been replaced by the Independent Evaluation Panel (IEP) (20);
- **Recipients of GHI funding** include health ministries (national or sub-national), United Nations (UN) agencies, international and local NGOs, CSOs, private sector (e.g. consultancy, digital start-ups, pharmaceutical), higher education institutions and research institutions. Many actors are keen to continue to receive funding from GHIs;
- **Donor agencies** (bilateral, multilateral and private foundations), which constitute the main funders of the GHIs;
- **Multilateral agencies** (such as WHO, other United Nations (UN) agencies, World Bank) and regional development banks, which work in the same field as the GHIs, often have country presence, and can act as collaborators or competitors (or hosts, in the case of the World Bank for the GFF).
- **Political and interest groups,** which exert pressure on donor governments and GHIs (lobby and campaigning groups, international NGOs, transnational corporations).

Historically there have been few incentives within any of the actors to maximise collaboration given the competitive funding landscape, but recently interactions between actors are becoming increasingly intricate, with some GHIs as central players (16) and growing inter-agency partnerships even between the GHIs. (21)

The types of power and influence wielded depends on the scope of the actor, which is summarised in Table 3 with reference to broad categories (acknowledging that there are nuances within each). Methods of wielding power are diverse, including funding power, influencing through formal governance structures like Boards, and normative power from organisations like WHO. The funders of GHIs were identified as the most powerful actors in the global analysis; they are the only actors that hold the ultimate sanction of withdrawing funding from the GHI ecosystem. The Boards were identified as the principal mechanism through which they can wield that power, but it was observed that this was not always exercised successfully. Reasons for this include that bilateral donors have diverse focal areas and tend to function in accordance with their own interests and values. This means that donor coordination and alignment can be weak. They are each accountable for their tax-payer-funded investments, hence they seek reassurance on fiduciary risks, as well as measurable impact. This also makes them attentive to the views of interest groups within their own countries. In addition, DAH departments within high income country (HIC) governments are required to be accountable to the wider foreign and economic policies and objectives of the country, and this creates additional layers of tensions and compromises for a purely health agenda. Some bilateral donors favour disease-specific investments, while others are more system-oriented. However, they too benefit from the GHIs as an efficient (for them) vehicle for aid spending. Some academic and CSO KIs perceived bilateral donors as prioritizing visible and rapid results to safeguard the health security of their own citizens, such as addressing infectious diseases and preventing their cross-border spread. Philanthropic foundations (which also fund GHIs) may have other interests, including using the GHIs as vehicles for projection of influence.

**Table 3.** Summary of interest and influence of major stakeholder groups.

| **Actors** | **Interest and position** | **Power and influence** |
| --- | --- | --- |
| GHIs | Interest in maintaining existence, which requires demonstration of results and being adaptable, expanding mandate where new needs are demonstrated.<br>Each GHI has its own incentives, which in funding GHIs are focused on fund flows and accountability for these. | Power formally sits with Boards, made up of diverse constituencies. However, not all constituencies are equally empowered or coordinated, leaving considerable influence in hands of senior leadership of GHIs.<br>Six-monthly meetings of a few hours cannot provide sufficient oversight so other modes of control slip in. |
| Bilateral funders | GHIs provide a useful platform for joint action for bilaterals, which are their major funders.<br>Each bilateral has to reflect domestic priorities but some (broadly, a European bloc, with | Considerable influence over GHIs as major funders (in proportion to contributions, broadly), however that influence is undermined by lack |
|  | others such as Japan and Canada) are more committed to integrated services and UHC, with higher risk tolerance to achieve more sustainable results. Whilst others (such as the US) are more committed to domestic political priorities, such as HIV, although this may be changing. | of coordination between them on reform agendas. |
| Multilateral organisations | Multilateral organisations play multiple roles in relation to GHIs, including:<br>- technical partners (e.g. through Accelerators, providing thematic coordination, and also through co-financing of programmes at country level, for example with the World Bank)<br>- rivals for bilateral and wider funding<br>- providing technical guidance to GHIs (e.g. WHO disease programmes and health system teams)<br>- grantees and implementing partners (e.g. UNDP)<br>- suppliers (e.g. UNICEF as a major purchaser of vaccines for Gavi)<br>Consequently, their interests are very mixed across the different organisations, as well as internally within each one | Influence at global level varies. A number, such as WHO, have normative power which affects the GHIs. Others are important as partners and implementers at country level. Some KI argued that the weakness of WHO was one of the factors in the large role of the GHIs. Many efforts have been made to coordinate this group with the GHIs, however, their influence is not strong enough to override internal incentives of GHIs. |
| Private foundations | Private foundations have contributed important sums to the GHIs, especially the Gates Foundation, which has invested in Gavi and the Global Fund in particular and is supportive of them, albeit sometimes as a 'critical friend'. | The Gates Foundation has significant influence through funding and board membership on some of the GHIs, while also supporting coordination mechanisms, such as the Accelerators. |
| Recipient government agencies (national and sub-national) | Government agencies have a broad interest in receiving financial, material and technical support from GHIs but there are diverse constituencies internally, with some stakeholders, such as disease programme directors and those represented on national GHI governance bodies, gaining resources and privileges (such as attending international meetings), while others with more integrated portfolios, such as planning, can find their jobs harder to do. | Recipient governments exercise power through their presence on the GHI Boards, as well as in local decision-making on grant applications etc. However, there was scepticism as to how formal board membership translated into real decision-making power by KIIs, due to informational barriers as well as the frequency and structure of meetings. Power in relation to grants was limited by bureaucratic requirements, though some countries have shown agility in making these work better for them.<br>Power dynamics on local governance bodies, such as the CCMs, will depend on the balance of constituencies and individuals (their interests, networks and capacities). |
| Non-governmental organisations, consultants and academics (local and international) | NGOs play diverse roles in relation to the GHIs, including as lobbyists, board members (representing civil society), sub-contracted consultants, and implementing partners. These create different positions.<br>- Some NGOs and consulting agencies, especially the large HIV-focused ones, have been strongly supportive of organisations like the Global Fund and resistant to reforms. Several universities in LMICs play the role of service providers and are powerful advocates of GHI funds.<br>- In the middle are some implementers and consultants, which may have critiques but are not able to voice them easily, due to their financial dependence.<br>- At the other end, are highly independent and hostile academics and CSOs which have highlighted the many problems created by the current operating modalities. | The major NGOs which can mobilise public pressure on funder governments and/or are represented on governing boards are reported to have considerable influence over the GHIs. Others (implementing NGOs, consultants and academics) have less influence on major issues, though they are engaged in technical consultations on more detailed areas, such as when organisational strategies are being revised. |
| Private sector providers and suppliers | The private sector has varied interests as it plays multiple roles in relation to the GHIs, including as supplier of inputs, partners in product development etc. | The private sector is often represented on GHI boards but does not feel very well engaged, according to our (limited number of) interviews. |

*Source: summarised by team based on analysis of KIIs*

### Source: summarised by team based on analysis of KIIs

Within the GHIs, senior leadership was seen as highly influential, not least because of the challenges noted for Boards (further discussed in the context section below). Technical power also sits with the GHI Secretariats, and especially the country grant managers (more so than technical advisory staff), who are in charge of fund disbursement, which is a key performance metric for GHIs, according to KIs.

> *“It’s the same program managers who developed the same applications or hired the same consultants to write the same applications. There are three-year time horizons, it’s short-term. Short-term money, short-term thinking and the grant managers…all of the incentives for the grant managers are to get the money out the door. That’s honestly the main key performance indicator: Get the money out the door.”* (Global KI)

The degree of financial dependency is a key variable in the position of national actors. In crisis-affected regions such as the Sahel, struggling with a reduction of domestic funding for health and the withdrawal of the main technical and financial partners, dependence on GHIs has increased and their support is highlighted as critical. (Southern and East Africa regional consultation KI).

Many of the actor groups, as noted in Table 2, have mixed positions and incentives because of the different roles they are playing and resources they may receive from the GHIs. The variation can be between departments within organisations as much as between organisations. Their power or influence is also varied. At country level, local NGOs were not reported to be influential on GHIs in general. South Africa presents a contrasting picture in that the Treatment Action Campaign was influential in improving access to prevention and treatment options for HIV in particular. (22) Globally, however the single interest lobby groups that campaign on certain health targets were viewed as highly influential in mobilising public opinion amongst voters and taxpayers. They can effectively bring pressure upon bilateral donors about how DAH budgets are allocated. This is reported by KI to be one reason why such a large proportion of the Global Fund’s budget (50%) is allocated to HIV.

> *“The epidemiology suggests that there should be more money for TB than HIV, and there’s no other money. It’s not like there’s another PEPFAR for TB.”* (Global KI)

The GHIs, by holding a significant portion of global health resources, have had an impact on the role of actors within some countries. This is particularly true for NGOs and some UN agencies. At the country level, some UN agencies and large NGOs are reliant on GHIs for “soft-funding” to pay key members of staff on their programmes. For instance, there has been a transformation of the UN from primarily a normative agency to a supplier and subcontractor, in many cases heavily dependent on GHI funding. The Pakistan case illustrates this phenomenon. Pakistan receives extensive funding for polio eradication and much of the effort is invested in eradication campaigns. UN agencies manage the campaigns, deploying a large number of staff and consultants supported by GHI project funding. However, government stakeholders are of the opinion that direct delivery campaigns, even if bringing good results, limit the development of country ownership and leadership (Pakistan KI). At the same time, some NGOs have also experienced a shift from advocating for health issues to assuming supply roles in response to the influence of GHIs.

WHO was often described by KIs at country level as weaker in its partner coordination role than desirable, absent from some of the roles perceived to be important parts of its function, and not managing to support UHC effectively. There are also potential conflicts of interests and inefficiencies as WHO applies to GHIs for funding from some country budgets, and also assumes the role of a supplier of both technical assistance and services in the presence of a weak government system. In all of these scenarios there is a risk that government systems are effectively bypassed and are not strengthened, with funding flows tilted more towards UN agencies and NGOs. Another key actor in several countries is The World Bank, in some cases providing its finance and convening power to bring bilateral funders and GHIs together for investment on specific country priorities.

Finally, more peripheral actors include the academic community, which is minimally involved in the implementation of GHI grants, though some evaluate their impact. They were amongst the most critical, highlighting problems with the whole current model of external aid and conflicts of interest in the aid landscape. This is also reflected in the literature which questions the role of “philanthrocapitalism”(23,24), use of for-profit consulting firms (25), and the pharmaceutical sector’s influence on GHIs.

A particular facet of the current complex global health funding environment around which there was considerable tension is the use of short-term consultants, particularly at country level where this is seen as boosting private interests and incomes over public service development (26) and again bypassing the strengthening of national health systems. Domestically there can be a revolving door of key, knowledgeable and highly skilled individuals between government, NGOs, GHIs and independent advisory work. They can also represent an unfortunate brain drain out of central government roles.

In addition, there can be a plethora of technical assistance both from the region and globally, often funded by GHIs or other partners, sometimes with unclear terms of reference, possibly overlapping activities and not aligned to country needs. The interests of international consultants versus local ones also emerged as a tension in all three country case studies.

> *“The Global Fund and other partners are helping Senegal to apply for grants and submit high-quality applications. Unicef, for example, recruits a consultant to support the country, notably at CCM level, as part of the elaboration of the GCS7. They have procedures, which require specific expertise, maintain the consultancy market and do not necessarily encourage local capacity building” (Senegal KI)*.

Some country KIs highlighted the way in which the complex systems operated by GHIs privilege experts and the disempowering effects this has on government staff.

> *“The experts are in charge and have taken total control of the organization. In some countries, 20 experts come and write a concept note … No concept note is written without experts.” (SEARO KI)*.

Health staff are another constituency, which often benefits from GHI funds in the form of per diems and salary supplements, which can however have very distorting effects on the health workforce (27–31). In-country health staff who are highly trained and knowledgeable about GHIs are sometimes recruited by the GHIs and assume roles as experts responsible for monitoring grant implementation, either in-country or at the GHI headquarters (Senegal KI). In South Africa, health staff are often recruited from the same geographical areas where GHIs support service delivery, and are paid higher salaries than those working within the public sector, leading to weaknesses within the system (South African KI)

Private sector KIs at global and country levels were willing to be more engaged with the GHIs but did not feel very much so at present.

> *“Engagement of private sector is important. All initial GHIs gave less importance to the private sector. The common notion was that private sector is not permanent and can go away. However, it is there to stay. Private sector and government sector are there to complement each other. Strengths of the private sector can better used to find an out of the box solution”* (Pakistan KI)

## Context

### Governance

The Boards of some of the GHIs were seen as innovative when first set up, with representatives from a range of constituencies, including implementing countries, donor countries, CSOs and the private sector. The GFATM’s Board has equal voting seats for donors and implementers, with 10 constituencies respectively. Within the 10 voting implementer constituencies, seven are implementer governments. Gavi also has representation from the vaccine industry and research and technical health institutes. Instead of a traditional board, the GFF established an Investors Group (32), which includes a range of actors, including UN agencies, recipient and donor governments, CSO, private sector, and youth representatives, and a Trust Fund Committee.

While the Boards of the GHIs are designed to monitor and ensure performance, there were varying perspectives on where the authority to challenge and rectify issues actually resided and how it was effectively exercised. Despite being theoretically representative, several KIs indicated that the Boards of some bigger GHIs have been structured in a way that fosters a balance of constituencies, resulting in rather slow and inefficient decision-making. Furthermore, KIs highlighted that the boards of GHIs can be very large and unwieldy, and this can also make consensus for change harder to reach. In addition, Boards can be at a disadvantage as Board members typically have short tenures, and this maintains an asymmetry in organisational knowledge and skills between the Boards and Secretariat, which has institutional memory.

In addition, KIs noted that there is a mismatch in the profiles of board members from the Global South and Global North, impacting their ability to effectively contribute and engage in decision-making processes. There are two key elements to this that came up in our interviews. The first is that the people sitting on Boards from the Global North are not of equivalent seniority to those representing the Global South - the example of government ministers representing the South whilst the North is represented by ‘bureaucrats’ from donor agencies was given. Second, the nature of the interaction appears to be unequal, with several KIs stating that it was not possible to “speak out” in Board meetings. Concerns were raised regarding the effectiveness of Board processes in facilitating active and open debates, especially for country representatives. It was observed that specific influential bilateral organisations, as well as certain large NGOs, hold more power than the recipient countries themselves. At county level, NGOs represented on boards may sometimes represent their own interests, more than those of the recipient communities (South African KI).

> *“On paper [GHI Boards are] diverse but I don’t think that the practical spaces that they provide actually allow people to speak in the way that they need to speak. It’s all muted and it all becomes politics and corridor speak. This is why I don’t go to [GHI] meetings anymore.”* (Global KI)

These “corridors” are shared by GHIs and bi/multilaterals in Geneva and Washington DC, but not with the Southern representatives, so it is more difficult for them to informally influence decision making. In addition, the lines of accountability are reported to be skewed towards funders, more than country health systems.

> *‘The accountabilities are to the capital donors and to getting the money out of the door. And there’s not enough accountability to real results in country or to efficiency-oriented concerns.*’ (Global KI)

The boards were also seen as not having the right technical expertise to address the challenges that the GHIs and the global health system now need to face, in particular those of strengthening health systems and achieving UHC.

> *“When you talk to [GFATM] about the importance of working with others to strengthen health systems in a way that’s not specific to HIV, you tend to get pretty blank looks… That’s not what they’re there for… They’re there to finish the job on HIV, and maybe TB and malaria.”* (Global KI)

Another aspect of unclear accountability at the global level was raised by some KIs in relation to the lack of transparency of reporting by some GHIs on their activities and investments as well as independent evaluations of their effect and cost efficiency.

Consequently, this fragmented funding landscape leads to the proliferation of plans, funds, reporting mechanisms, and auditing processes. Such fragmentation not only contributes to inefficiency but also proves to be ineffective, overwhelming the capacity of the recipient country to effectively manage these resources.

> *“You know there’s multiple reporting channels, there’s multiple. And so it’s a complete nightmare (South African KI)*

> *“Gavi has its immunisation financing, technical support and then polio has its polio transition. And GFF has its UHC alignment. And we’re just all pulling the same people to the same meetings. And the organisations themselves aren’t accountable for the fact we just distract and are selling our own products and justifying our own existence through these processes.”* (Global KI)

Governance challenges were highlighted in the case studies - for example, in Senegal, where the presence of multiple governance structures across GHIs generates high transaction costs and risks of uncoordinated initiatives for the government (120) (see also Boxes 1-3). Each GHI has its own operating methods, procedures, contracts and coordinating bodies.

In the case of the GFATM’s Country Coordinating Mechanism (CCM), some concerns regarding its current make-up and operations were also raised, as it is typically representative of specific interest groups who may also be funding recipients, aligned to the three diseases, while they may lack the technical expertise needed to develop strong health system strengthening (HSS) proposals. Other concerns relate to the possible blurring of roles and responsibilities, and potential conflicts of interest. For example, in South Africa, the Department of Health is both a member of the CCM and a principal recipient. Furthermore, the South African National AIDS Council (SANAC) runs the CCM, which is positively viewed by some as indicating local leadership. SANAC is however also a recipient of GFATM money and implements programmes within health facilities. The Secretariat for SANAC is also the Secretariat of the GFATM. There is however strong CSO representation and SANAC is co-chaired by the country’s deputy President (33).

New institutional interests can also be set up as a result of siloed planning and funding:

> *“The Global Fund model and the Gavi models are interesting. They say they are not going to establish their own in-country presence, but what they’ve done is create their own in-country institutional monsters in some respects. We have the ministries of AIDS, right*?” (Global KI)

At country level, accountability to GHIs (focused on managing financial risks) can take precedence over accountability to communities and national entities (for performance).

> *“Within the countries we lose a lot of efficiency because the country teams have to set up no objection procedures, the fiduciary agencies have to validate the implementation, we lose efficiency. Implementers spend more time looking for ways to comply with FM [financial management] directives… regard is more focused on satisfying Geneva than communities”* (SEARO KI)

Other concerns included that reports are sent to ‘Geneva’ or to GHIs’ funders or stakeholders, but not necessarily to the local policy-makers responsible for delivering health services (Addis consultative meeting KI). Multiple KIs urged better country engagement and transparency regarding funding to enable collaborative action plans.

> *“From a country perspective, I would give them 4/10 for improving health outcomes; 2/10 for improving the health system capacity, 1/10 for graduating from dependence on international finance, and 0/10 for ownership by the government and supporting their policies.”* (Global KI)

### Financing

In a context of plateauing DAH (34), the overall environment is marked by competition between GHI actors for funds, which drives expanding mandates to ensure continued relevance, for example in the face of new threats such as COVID-19 – counterbalanced by long-standing initiatives to improve alignment between GHIs (Figure 2).

**Figure 2.**
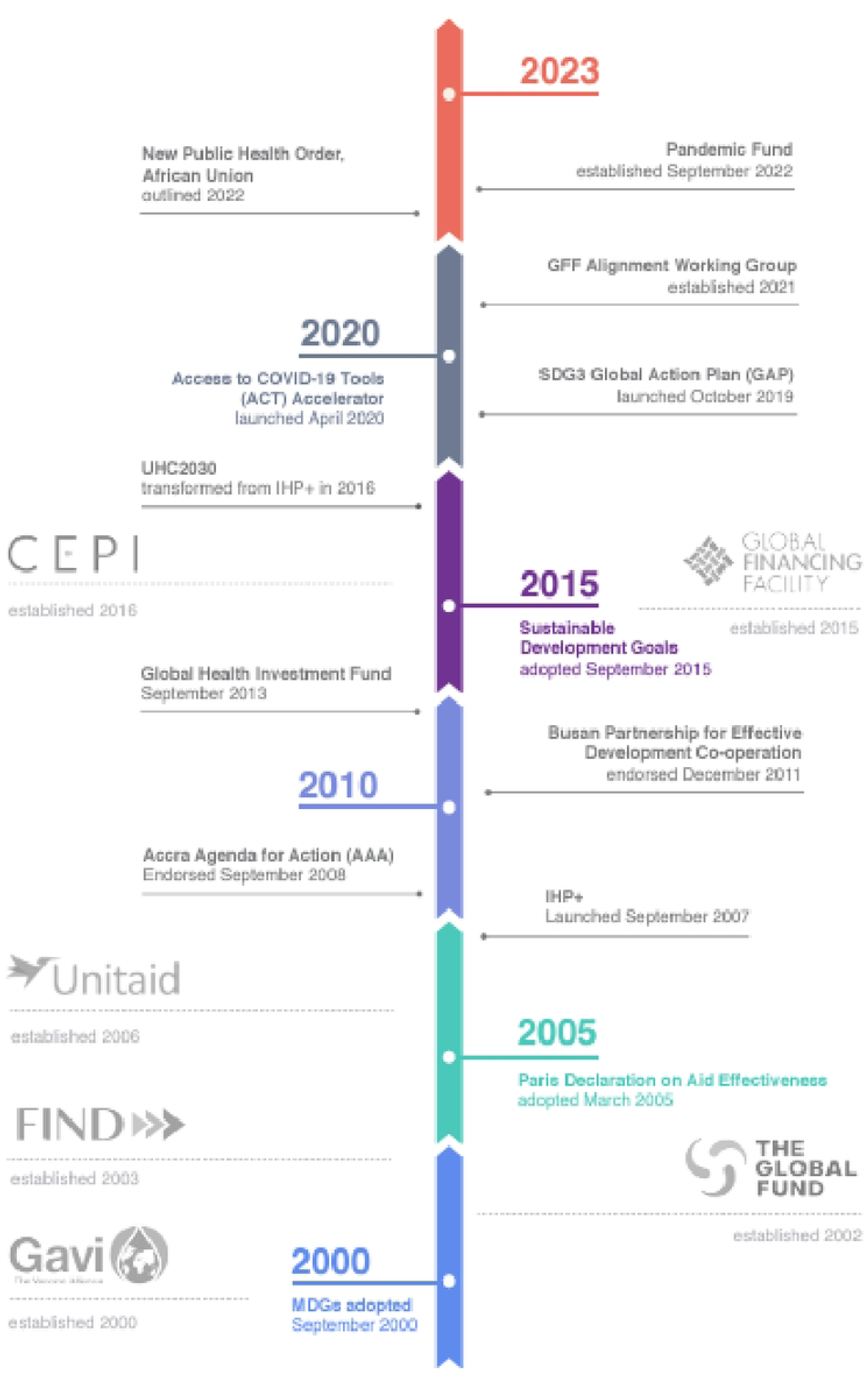
Creation of GHIs and some alignment initiatives, 2000-2023. **Source**: Witter et al. 2023 (**4). Image credit: Claudia Molina**

Global KIs perceived competition for funding between GHIs and other global-level organisations, creating a sense of a zero-sum game, where funds may also not align with the actual needs in terms of disease burden or the functional role of different organisations. The competition for funding from the same pot of money was perceived to be likely to contribute to a perceived eagerness of GHIs to take on new roles and expand their mandate, as organisations jostle for roles and funding. The existing system of staggered replenishments by GHIs was perceived as challenging for bilateral donors and governments of LMICs to manage (35–37) and there were concerns regarding the overall financial sustainability of the repeated, increasing GHI requests for replenishment.

At country level, dependence on GHI resources can lead to imbalances in relation to priority areas and loss of alignment. In Senegal, for example, despite low prevalence, HIV programmes continue to receive substantial funding, whereas non-communicable diseases, which are more prevalent, lack sufficient resources (KII and (38)). This was echoed in the South African case study, where despite the high HIV prevalence concerns were raised that not enough finances were being directed to non-communicable diseases and strengthening of primary health care.

At the country level, some GHIs wield considerable power, depending on their contribution to the country’s domestic funding. GFATM and Gavi are important funders to governments, NGOs and civil society. A comparison of WHO’s Global Health Expenditure Database (April 2023 update) (39)and OECD Creditor Reporting System (40) data indicates that Gavi and GFATM gross disbursements accounted for a larger combined budget than domestic government funding in seven sub-Saharan African countries^2^ in 2020, giving these two institutions considerable influence. As an interesting contrast, in South Africa DAH constitutes less than 5% of total health expenditure, with the GFTAM providing the largest share of funding for HIV and to a lesser degree TB and malaria.(39) KIs reported that this small contribution to the overall budget does limit their power at governmental level. As in other countries, GFATM and Gavi also work through a variety of channels and by empowering non-state actors or disease-specific programmes they are still capable of creating advocates for them. Lack of transparency can also cause challenges for managers at devolved levels:

> *“In Ghana, in talking to district managers, they were so frustrated because these donors were coming in, running their funding off budget and basically bypassing them… The district managers have very little power in how these resources are allocated, but they’re held accountable for delivering within their districts. It’s crazy, right? And there’s so much frustration at that level. I think from a governance side they should be very transparent.”* (Global KI)

There are also imbalances within government, in that funds go disproportionately to some programmes (such as HIV/AIDS and malaria), which creates inequities and also vested interests amongst some Ministry departments. For instance, in Mozambique, a KI reported that 80% of the funding received is for HIV, which creates a set of vested interests out of balance with the rest of the health system, and little incentive for these recipients to support a more integrated system. The ability to gain such disproportionate benefits from GHI funding, including as a result of the opaque mapping of funding to public expenditure, creates pockets of strong resistance to reforming the GHIs as they are currently functioning at country level.

By contrast, GFF works through more a integrated funding mechanism, which raise different concerns about fungibility.

> *“Financing takes the form of budgetary support or trust funds, producing a substitution effect between donors and governments. How can we explain the fact that while budget support is increasing when, health expenditure and needs are not being covered?”* (SEARO consultation KI)

Moreover, provision of funding is perceived as not tied to a country plan led and owned by Ministers of Health and instead is tied to programmatic funding cycles of Gavi and GFATM, with an imperative to disburse funds rather than support national planning. This results in the provision of fragmented ad hoc funding and exacerbates frustration within country governments at being powerless to direct funding or ensure accountability:

> *“The power lies with GHIs so far. They send you the support but you do not have a say. If you do not have a say, you do not have power”* (Pakistan KI; see also Boxes 1-3)

#### Box 1. Country Case Study: Pakistan

Donor financing in Pakistan, inclusive of bilateral agencies, multilaterals and GHIs, has typically been less than 2 percent of the total health expenditure (74,75). Gavi finances vaccines, cold chain, advocacy and community outreach support for immunization-Polio eradication. GFATM extends the largest support to TB diagnostics, which includes integration of the private sector. It also makes contributions towards malaria control and HIV prevention through communitybased outreach information systems strengthening, and awareness(76). Global Financing Facility has recently started contributing to Pakistan and will be contributing to maternal care as part of pooled financing with the World Bank (77)

**Challenges**

**Governance, coordination and alignment**

- Competing technical assistance plans between donor agencies and GHIs, and between government and GHIs, resulting in duplication of assistance and divergent priorities
- Weak country capacity for aid coordination, realistic target setting and planning but little investment in capacity building
- Lack of coordination between federal and provincial governments, exacerbated by fragmented projectized funding by GHIs, constrains cohesive country planning
- Leadership erosion with frequent leadership changes of health secretaries and disease managers

**Health Systems Strengthening and integration**

- Several ongoing local health reforms but GHI funding not integrated into reform planning, hence constraining cohesion and sustainability
- Uneven capacity of disease planners and health system managers
- GHI prioritization of disease control programmes is insufficiently backed with local health systems strengthening support
- Disease control efforts are not framed within the larger ambit of Primary Health Care
- Large private sector but not effectively harnessed for disease control and PHC

**GHI financing**

- Funding and disbursement is driven by donor-led burden of disease analysis with less consideration of local health systems realities.
- Ad hoc use of external finance as standalone projects rather than integration into ongoing initiatives for sustainability and efficiency
- Multiple parallel funding streams by GHI constrains oversight and coordination of external financing
- World Bank aspirations to leadership under pooled funding but lack of integration of lessons learned from past attempts at pooled funding

**Monitoring and performance accountability**

- Overambitious targets set by GHIs, not contextualised to local health systems realities and opportunities within existing reforms
- Low political voice of the government to articulate accountability needs as well as weak systems and staffing limits accountability and counter correction measures.
- Accountability constrained by lack of a central accessible repository of funding / projects data

#### Box 2. Country Case Study: South Africa

GHIs contribute less than 5% towards health financing in South Africa. PEPFAR and GFATM are the largest donors. FIND, Unitaid and CEPI fund non-state actors. Gavi and GFF have no in-country presence. South Africa is a donor to Gavi and GFATM.

**Challenges**

**Governance, coordination, and alignment**

- Lack of in-country alignment of GHIs’ priorities and activities with country health plans and priorities
- Separate in-country GHI coordination and resource mobilisation mechanisms
- GHIs tend to by-pass government structures and directly fund non-state actors
- Civil society not sufficiently active or strong to hold GHIs accountable for in-country activities

**Health Systems Strengthening and integration**

- Prioritized disease control programme by GHIs; lack of support for local health systems
- strengthening (HSS) (e.g. Universal Health Care) reforms, resulting in fragmentation
- Duplication of systems (information, health financing, etc) resulting in increased burden on health managers
- Bypassing of local experts in favour of international ones who do not understand the local contexts

**GHI financing**

- Funding in areas of donor interest with little consideration of local health systems realities.
- Funding for implementation not always strategic or sustainable (e.g. use of funds for specific line items/activities)
- Bypassing of national government financing system/lack of reporting transparency; therefore, government cannot account for all GHI funding
- Donor funding tend to have conditionalities or restrictions attached to them which may be at odds with country priorities

**Monitoring and performance accountability**

- No formal in-country governance or accountability mechanisms that mandate that GHIs first report findings and challenges to country before reporting to their stakeholders (e.g. Boards)
- Limited evidence of the real effect or impact of GHIs on health outcomes or whole-system effects.
- Large GHI datasets and multiple reporting systems undermines the country’s health information system processes; insufficient coordination, integration/alignment thereof

#### Box 3. Country Case Study: Senegal

According to the most recent National Health Accounts (NHA 2017-2021), donors finance almost as much as the state (22.7% vs. 25.7%) for all health expenditure, while households support 43.5%. (38) However, the Senegalese government finances less than 10% of healthcare expenditure for the three GFATM diseases. For malaria, USAID is also heavily involved in funding. Under the Global Financing Facility (GFF) investment plan, the government of Senegal was expected to contributed 34% of funding by the end of 2021 (78).

**Challenges**

**Governance, coordination and alignment**

- Lack of communication and coordination between the GHIs in Senegal
- Lack of comprehensive understanding of the overall landscape of GHIs by national stakeholders,
- National experts leave the civil service to become consultants to GHIs
- Coordination bodies and platforms are not dynamic and effective (“lethargy”)
- Global actors are far from the real world and population needs/lives
- Power imbalance in term of establishment of priorities
- Language barriers (almost exclusive use of English)

**Health Systems Strengthening and integration**

- Fragmentation of initiatives; program verticalization
- Implementation gap (delays in implementation of interventions)
- Insufficient investment and impact of GHIs on health system strengthening (HSS), despite recent efforts
- Investments in specific diseases inadequately benefit the broader healthcare system

**GHI financing**

- Cumbersome procedures
- Multiplicity of windows, interlocutors, and methods of financing
- Funding spread over activities instead of building sustainability

Some countries have shown notable progress in adopting a more integrated approach – for example, Malawi is currently making progress on greater integration (41); additionally, Ethiopia, Rwanda, Somalia, and certain provinces of South Africa have been recognised as enforcing a more harmonised approach across funders, including GHIs (42). There is scope for countries to shape GHI support, where will and capacity exist, but this is not always facilitated by the GHI requirements.

According to South African KIs, GHIs and larger donors often by-pass government, due to lack of trust in government, and provide direct funding to NGOs, CSOs, Parliament and higher education and research institutions, undermining control and overview of central institutions such as the Department of Health and Treasury. Reportedly, approximately half of the GFATM funds are allocated to government recipients, but even among those, a significant portion remains off-budget (40,43). In pursuit of their goal to channel 55% of funding through government systems by the end of 2021, Gavi has made strides in increasing the share. However, as of 2021, only 41% of the (non-commodity) funding had been directed through these systems.

Country KIs are also sceptical about the small proportion of funding that is expended within countries. Only operational funds of country grants are actually spent in the country whereas the bulk of the funding often comprises supplies which are internationally procured as local vendors are not pre-qualified for GHI procurement. There have been long-standing concerns of lack of international community support to boost the local industry for supplies production, which leads to a cycle of dependency on GHI funding.

> “*Local vendors are not pre-qualified. Therefore, we send back 70% of the funding to the donors through international procurement and that at a very high cost compared to the local purchase”*. – (Pakistan KI)

Despite the focus on minimising fiduciary risks, there are also concerns that the GHIs (GFATM and Gavi in particular) may inadvertently contribute to or escalate corruption risks. This concern stems from the use of multiple independent bank accounts and off-budget systems, which can create opportunities for financial irregularities. Periodic crises have been linked to poor accounting practices and inadequate tracking of fund usage (44–48).

## Narratives and framing

### Performance narratives

GHIs justify themselves in relation to results in their focal areas, but there is much contestation about how those results are generated and whether they reflect others’ investments in the results chain. While the GHIs are recognised to have made substantial contributions to the results chain for their focal areas, many global KIs and the literature (49–51) reported that some of them over- claim results, especially on blunt indicators such as ‘lives saved’. Specifically, they are perceived to claim credit for the entire outcome of broader investments, which encompassed contributions from LMIC governments and from other funders. In some cases, reported results have been primarily based on modelling, rather than comprehensive evaluations.

> *“They get the receipts [for inputs], but they don’t really know what they are producing.”* (Global KI)

The GFF has moved away from this model and reports on assessed contribution to national/country results, with a clear line of sight to the nature and value add of the GFF contributions, which made their reported results less questioned by KIs. However, this was mentioned by some KIs as having weakened their case for impact in comparison to some other GHI claims. This shows the pressure that GHIs are under to compete and ‘out claim’ one another in order to attract or maintain funding.

In response to concerns about health system impacts (52,53), there has been an increased focus in GHI policies on ‘HSS’ investments. However, with GFATM the classification of spending as supporting resilient and sustainable systems for health (RSSH) was also questioned by global KIs, who claim that what is counted as RSSH and what is seen as disease-specific does not follow a clear logic. There has been ongoing debate and lack of clarity around how much money spent by GFATM and Gavi can be classified as actually strengthening the health system in a sustainable way (54). Various attempts to classify expenditure have been made, ranging from 27% to 7% of investment (55,56).

Several KIs mentioned that the narrative is dominated by what they interpreted as powerful and vocal interests grouped around the GHIs at global level, which have strong interests in emphasising the strengths and successes of GHI activities, and have the resources to amplify this message. This is in contrast to more critical voices at country level and globally, which are not able to project their views with such power. As was highlighted in the governance section, some Board members also feel less able to speak out in the face of these power differentials.

### Narratives about capacity

At the national level, particularly in contexts of financial dependence, there can be a mutual blame game, in which GHIs and other partners lament lack of national capacity and planning which forces them to play a dominant role, while national counterparts resent their lack of control, ownership and independence, blaming GHIs for undermining these and not building their capacity. Both sides have an element of justice and the behaviour on both sides can reinforce continued patterns of this nature.

> *‘The government is meant to set targets but GHIs set priorities because the government is unable to define priorities. The country is thus pushed to achieve targets set elsewhere with the local context (e.g. economic climate, available resources, burden of disease, political realities) ignored. This is because of very limited state capacities that is reflected in a weak national programme, a Health Department with no vision or capacity, the absence of a public health approach, (realistic) health financing strategy or medium-term (five-year) and long-term (15-20 year) plans.”* (Pakistan KI)

Part of the challenge relates to the timeframe and institutional incentives of GHIs, which have relatively short funding cycles, while building capacity takes longer and is harder to measure.

> “*[GHIs are] top-down, selective, short-termist, and kind of have a bias towards delivering things that can be measured. In a neglect of important things that need to be improved or strengthened. But which can’t necessarily be measured in a way these initiatives tend to want to measure things – which is by counting things.”* (Global KI)

> *“So health systems work is by nature difficult. Part of what it achieves is preventing more bad things from happening. That’s always difficult to gauge and assess”* (Global KI)

Some of the divergence of discourse on the impact of GHIs relates to respondents focusing on different outcomes – in particular, short-term gains in coverage in specific areas versus longer term changes to how system operate. The fact that GHIs primarily fund inputs means that there is continuing dependence in the longer term.

> *“We’ve done really well over 20 years in bringing down the incidence rate of HIV, saving people from dying of HIV with TB and malaria as well. But of course as soon as the money dries up, that all starts to disappear, all those gains, and that’s what we saw over COVID, right?”* (Global KI)

### Narratives about risks

It is also important how risks are framed. The GHI systems are in many cases primarily designed to prioritise minimising fiduciary risk, which is crucial for donors. However, that may not be inherently more important than addressing programme and system risks, such as the risk of failing to achieve progress, failing to strengthen programmes, or causing unintended harm to health systems. Enhancing effectiveness may involve increasing flexibility, even if it results in higher fiduciary risk. This aspect becomes particularly significant in FCAS, where the circumstances are dynamic and require adaptability. KIs point out that more work needs to be done on balancing the costs of different approaches and using more context-adapted measures.

> *“There is a problem with the financing flexibility. The Global Fund, for example, has very strict budget lines and in conflict settings, it does not allow us to adapt according to the current situation.”* (EMRO consultation KI)

### Narratives about potential reforms

The data revealed divergent perspectives on the role and possible future path of the GHIs (see Box 4). Some implementers and funders were incrementalist in their approach to change, whereas other country-level actors, multilaterals, and academics tended to be more radical. There is also a lot of variation within these groups. It is notable that there were surprisingly critical voices from within the GHIs themselves, reflecting the divergent pressures that staff within them are having to manage.

#### Box 4. Reform scenarios and narratives

Three predominant reform narratives emerged from the interviews and consultations. These are summarised here.

1. **Narrative of status quo** – this narrative, predominantly emanating from some parts of the GHIs but also some of their implementing partners, focuses on the big benefits delivered by GHIs; it views the GHIs as one of the more adaptive, successful elements of the global health infrastructure (‘why are you picking on us?’), on their successful mobilisation of funds (with a threat of their withdrawal if GHIs were too radically altered), their focus on vulnerable populations and innovative models of governance and financing. Problems that have arisen are presented as largely due to weaknesses of systems and capacity at country level. The GHIs should therefore continue to operate broadly as they do, with minor adjustments.
2. **Narratives of radical reform** – this narrative, which emanates from a range of respondents (academics, partners in multilaterals, also some GHI staff) highlights that GHIs have been overselling their success, as well as (in some cases) causing harms through fragmented, distortionary funding, and not focusing on the need to build sustainable, integrated systems. Further, they offer poor efficiency through input financing, are prolonging their own mandates beyond the original planned timespans, have low accountability to beneficiary governments, lack transparency on data, and have imposed high costs for governments and others to access grants though complexity and lack of coordination between GHIs and other actors. An end date should be set for the GHIs, either very soon or in the foreseeable future.
3. **Narrative of moderate/iterative reform** – in this view, which emanated from a range of respondents including country partners and funders, these GHIs do make an important contribution but their systems need to evolve to focus more on transition, capacity building, sustainability at country level, alongside the provision of global public goods, with recognition of the ongoing financial dependence for a smaller group of countries which are low income and/or fragile and conflict-affected. The focus of reforms should be on improving the functioning of the GHIs, which could include a range of actions from merger to shared functions, better alignment with country systems and one another, changed processes to reduce transaction costs for governments and implementers, and more support for integrated health systems.

The positive narrative about results noted above makes changes to the status quo more difficult. GHIs rely heavily on these narratives to make the case for their continued importance and existence, providing information systems and data to support their positions. At the same time, critical narratives emerged from our interviews, which support radical reforms. There is a discrepancy between these more radical voices and the official narratives within GHIs about reform, which weakens the possibility of agreement on the way forward.

The positive narrative about results noted above makes changes to the status quo more difficult. GHIs heavily rely on these narratives to make the case for their existence, and also use information systems and data to support it. At the same time, critical narratives emerged from our interviews, which support radical reforms. However, these narratives are not the official GHIs ones and there is a discrepancy between the official and informal narratives within GHIs about reform, which weakens the possibility of agreement on the way forward.

While reforming existing institutions is challenging, establishing new institutions appears to be an altogether easier route to plan to respond to new global challenges. Hence proliferation and fragmentation are perpetuated, impacting on recipient countries. Over the past few years, several new global funds have been created, including the Global Oxygen Alliance (57), the Hepatitis Fund (58), Health4Life Fund (59), the Pandemic Fund (60), and the Health Impact Investment Platform (61). The relevance, functioning and unintended consequences of these new funds, largely supported by the same bilateral donors, UN agencies and foundations, need to be evaluated. They add a new layer of complexity and fragmentation to the global health architecture and at national level, where each initiative focuses on a specific field, such as sexual and reproductive health and rights, HIV, or innovation, and operates with its own programs, governance structures, mechanisms, and approaches.

> *“The mechanisms are fragmented, but the public health problems they tackle are not”* (Senegal KI)

Another potential reform that was mentioned is the expansion of mandates of existing GHIs. However, some interviewees, especially global KIs, expressed concern about what they perceived as constantly expanding mandates, particularly regarding the GFATM and Gavi. They pointed out that these organisations have been expanding their roles and venturing into new areas, such as HSS (52,56). However, in their opinion, there is little evidence to suggest that GHIs are appropriately structured and technically equipped to handle these responsibilities effectively (South Africa KI; regional consultation).

## Discussion

In this article, we examined the role of GHIs within the global and national health architecture from a political economy perspective in order to understand patterns, points of resistance and possibility for reforms. This work is original in that there have been many analyses of and critiques of the GHIs but none which have looked with the lens of political economy, bringing in views from a large range of global, regional and national experts.

The current arrangement, with its strengths and weaknesses, is not accidental but emerged from a specific period which focused on reaching global goals on priority diseases, especially communicable ones (62), and when international funding was growing. Since then, the landscape has changed, particularly in relation to the emergence of non-communicable diseases and the health impacts of climate change, and financing for international support is under strain. However, the structures which were established 20 years ago have created a path dependency, with large, complex bureaucracies (in some cases; the scale is very varied across them) which have momentum and can resist reforms, as well as a large network of clients (including governments, implementers, consultants, etc.) who are interested in maintaining the status-quo.

Reflecting on the lessons that KIs and literature highlighted in relation to previous efforts at coordination and alignment, it is clear that individuals and organisations follow their own incentives, which need to be altered for behaviour change to follow. Voluntarist approaches to reform, which do not change rewards and sanctions are unlikely to gain traction (63,64).

The actors involved are numerous, diverse, interconnected and have interests which largely favour status quo or at most incremental reforms. These actors do not fit into neat categories and even at individual level can play multiple roles – for example, benefiting from being a consultant to GHIs at national level, while also holding a more critical perspective in a government role.

The GHIs themselves are also part of a wider network of DAH organisations, which interact with GHIs, with country health systems and with one another to influence outcomes, which makes reform highly complex. All are responsible and none are, which is a perfect setting for mutual blame and inaction on change.

Power to bring about change is not evenly distributed – some actors have more power and influence, especially major funders and senior leadership in the GHIs, but they also have to create consensus, work in coordinated ways and draw on wider legitimacy if they wish to enact reforms. For that process, which started with the Lusaka Agenda, the ability to draw on powerful narratives and clear accountability measures for reform will be significant.(65) Ultimately, all the elements of the political economy framework emerged as important here: the position and power of key actors, but also the context factors (financing flows and governance structures) which affect how GHIs function and how decisions are made, and the narratives and framing which influence both whether change is seen to be needed and what form it might take.

It is important to restate the differences between GHIs and note that the three country-facing GHIs exist on a continuum of integration with national systems, with the GFF most integrated through its provision of public financing, while Gavi is able to pool funds at national level and the Global Fund is least enabled to operate in that way. At national level, there are also variations in the dynamics observed in this study; for example, countries with greater financial dependence on the support of GHIs typically raised more concerns about their functioning, while better funded health systems (or sub-national components of health systems) were better able to use GHI support in ways that did not disrupt their operations.

As the GHIs continue to evolve in a dynamic global health environment, the deployment of political economy as a lens to understand what is possible, to understand change and its absence, and to strategise around building coalitions for reform (9) will continue to be very relevant from both an academic and policy perspective. (66)

## Conclusion

This paper has highlighted some of the key critiques and current dissatisfactions at national level with GHIs that are most active within country health systems. It has also described how the GHIs are part of a wider complex and interdependent ecosystem and that their role has evolved in relation to other actors, all of which play a part in the patterns noted here. Reform of the GHIs will involve changes by these wider actors, especially the funders, recipient countries, senior leaders in GHIs and influential NGOs, and will reflect shifting interests and narratives. Potential for change comes from the current perceptions of constrained resources and increasing threats, but this does not guarantee strengthening of the role of GHIs unless consensus is reached around narratives of how the current system is working and options developed which serve the interests of key constituencies. Political economy analysis can help to highlight these issues and point to strategies for managing them.

## Data Availability

All data is contained in the article and supporting references and sites

https://futureofghis.org/research-other-inputs/reimagining-the-future-of-global-health-initiatives-study/

## Acknowledgements

We would like to acknowledge the support of the Government of Norway and the Wellcome Trust in funding and guiding the original research. The opinions expressed here are however the responsibility of the authors alone. We would also like to thank all of our many key informants for their insights, and thank ReBUILD for Resilience research consortium for supporting publication of this article.

## Footnotes

1 By taking the last replenishment total and dividing by the three-year cycle; not a measure of actual expenditure per year

2 Central African Republic, Democratic Republic of the Congo, Eritrea, Guinea, South Sudan, Uganda, Zimbabwe

## References

1. Institute For Health Metrics and Evaluation (IHME). Financing Global Health [Internet]. Seattle, WA; 2024 [cited 2024 Aug 10]. Available from: http://vizhub.healthdata.org/fgh/%E2%80%8B

2. World Bank Group Development Finance. Understanding Trends in Proliferation and Fragmentation for Aid Effectiveness during Crises [Internet]. 2022 Jul [cited 2024 Aug 10]. Available from: https://thedocs.worldbank.org/en/doc/ef73fb3d1d33e3bf0e2c23bdf49b4907-0060012022/understanding-trends-in-proliferation-and-fragmentation-for-aid-effectiveness-during-crises?_gl=1*d1g7ww*_gcl_au*MjY4MjU3MjgxLjE3MjIzMTk1MzU.

3. Mwangangi M, Røttingen JA. The Lusaka Agenda: Conclusions of the Future of Global Health Initiatives Process [Internet]. Lusaka; 2023 Dec [cited 2024 Aug 10]. Available from: https://futureofghis.org/final-outputs/lusaka-agenda/

4. Witter S, Palmer N, James R, Zaidi S, Carillon S, English R, et al. Reimagining the Future of Global Health Initiatives, Research Report [Internet]. 2023 [cited 2024 Aug 10]. Available from: https://futureofghis.org/research-other-inputs/reimagining-the-future-of-global-health-initiatives-study/

5. Sundewall J, McCoy D, Brugha R. Correspondence: Global health initiatives and country health systems. The Lancet. 2009 Oct 10;374:1237–8.

6. Cohn J, Russell A, Baker B, Kayongo A, Wanjiku E, Davis P. Using global health initiatives to strengthen health systems: a civil society perspective. Glob Public Health. 2011;6(7):687–702.

7. Hanefeld J. How have global health initiatives impacted on health equity? Promot Educ. 2008;15(1):19–23.

8. Zicker F, Faid M, Reeder J, Aslanyan G. Building coherence and synergy among global health initiatives. Health Res Policy Syst. 2015;13:1–8.

9. Witter S, Sparkes S, Bertone MP, Ozcelik E. Political economy analysis for health financing: A ‘how to’guide. World Health Organization; 2024.

10. Fritz V, Levy B, Ort R. Problem-driven political economy analysis: The World Bank’s experience. World Bank Publications; 2014.

11. Bertone MP, Wurie H, Samai M, Witter S. The bumpy trajectory of performance-based financing for healthcare in Sierra Leone: agency, structure and frames shaping the policy process. Global Health. 2018;14:1–15.

12. Harris D. Applied political economy analysis. A problem-driven framework London: Overseas Development Institute. 2013;4.

13. Varvasovszky Z, Brugha R. A stakeholder analysis. Health Policy Plan. 2000;15(3):338–45.

14. Balane MA, Palafox B, Palileo-Villanueva LM, McKee M, Balabanova D. Enhancing the use of stakeholder analysis for policy implementation research: towards a novel framing and operationalised measures. BMJ Glob Health. 2020;5(11):e002661.

15. Schmeer K. Stakeholder analysis guidelines. Policy toolkit for strengthening health sector reform. 1999;1:1–35.

16. Hoffman SJ, Cole CB. Defining the global health system and systematically mapping its network of actors. Global Health. 2018;14:1–19.

17. Fergus CA. Power across the global health landscape: a network analysis of development assistance 1990–2015. Health Policy Plan. 2022;37(6):779–90.

18. Victoria Fan and Eleni Smitham. Center For Global Development (blog post). 2023 [cited 2024 Aug 10]. Is the New Pandemic Fund Where Vertical Can Finally Meet Horizontal? Available from: https://www.cgdev.org/blog/world-banks-new-pandemic-fund-where-vertical-can-finally-meet-horizontal

19. Micah AE, Bhangdia K, Cogswell IE, Lasher D, Lidral-Porter B, Maddison ER, et al. Global investments in pandemic preparedness and COVID-19: development assistance and domestic spending on health between 1990 and 2026. Lancet Glob Health. 2023;11(3):e385–413.

20. Office of the Inspector General. The Global Fund. 2021 [cited 2024 Aug 10]. Evolving the Technical Review Panel Model. Available from: https://www.theglobalfund.org/en/oig/updates/2021-11-04-evolving-the-technical-review-panel-model/

21. de Bengy Puyvallée A. The rising authority and agency of public–private partnerships in global health governance. Policy Soc. 2024;43(1):25–40.

22. Heywood M. South Africa’s treatment action campaign: combining law and social mobilization to realize the right to health. J Hum Rights Pract. 2009;1(1):14–36.

23. Birn AE. Philanthrocapitalism, past and present: The Rockefeller Foundation, the Gates Foundation, and the setting (s) of the international/global health agenda. Hypothesis. 2014;12(1):e8.

24. McGoey L. Philanthrocapitalism and its critics. Poetics. 2012;40(2):185–99.

25. Eckl J, Hanrieder T. The political economy of consulting firms in reform processes: the case of the World Health Organization. Rev Int Polit Econ. 2023;30(6):2309–32.

26. Eckl J, Hanrieder T. The political economy of consulting firms in reform processes: the case of the World Health Organization. Rev Int Polit Econ. 2023;30(6):2309–32.

27. Hanefeld J. The global fund to fight AIDS, tuberculosis and malaria: 10 years on. Clinical medicine. 2014;14(1):54–7.

28. Craveiro I, Dussault G. The impact of global health initiatives on the health system in Angola. Glob Public Health. 2016;11(4):475–95.

29. Calain P, Abu Sa’Da C. Coincident polio and Ebola crises expose similar fault lines in the current global health regime. Confl Health. 2015;9:1–7.

30. Ooms G, Van Damme W, Baker BK, Zeitz P, Schrecker T. The’diagonal’approach to Global Fund financing: a cure for the broader malaise of health systems? Global Health. 2008;4:1–7.

31. Spicer N, Walsh A. 10 best resources on… the current effects of global health initiatives on country health systems. Health Policy Plan. 2012;27(3):265–9.

32. Global Financing Facility (GFF). Global Financing Facility. 2024 [cited 2024 Aug 10]. Investors Group (webpage). Available from: www.globalfinancingfacility.org/governance/investors-group

33. English R, Dudley L, Daniels K, Hendricks L, Blows S, Pillay Y. Reimagining the Future of Global Health Initiatives study: South Africa country case study [Internet]. 2023 [cited 2024 Aug 25]. Available from: https://d2nhv1us8wflpq.cloudfront.net/prod/uploads/2023/08/Appendix-9-South-Africa-country-case-study.pdf

34. Chang AY, Cowling K, Micah AE, Chapin A, Chen CS, Ikilezi G, et al. Past, present, and future of global health financing: a review of development assistance, government, out-of-pocket, and other private spending on health for 195 countries, 1995–2050. The Lancet. 2019;393(10187):2233–60.

35. Clinton C, Sridhar D. Who pays for cooperation in global health? A comparative analysis of WHO, the World Bank, the Global Fund to Fight HIV/AIDS, Tuberculosis and Malaria, and Gavi, the Vaccine Alliance. The Lancet. 2017;390(10091):324–32.

36. Sridhar D, Woods N. Trojan multilateralism: global cooperation in health. Glob Policy. 2013;4(4):325–35.

37. The Lancet. The Global Fund: replenishment and future-proofing. The Lancet. 2022 Sep;400(10355):787.

38. Ministère de la Santé et de l’Action Sociale (République du Senégal). Rapport des comptes de la santé 2017/2021 [Internet]. Dakar; 2022 Dec [cited 2024 Sep 28]. Available from: https://p4h.world/app/uploads/2024/01/RAPPORT-COMPTES-DE-LA-SANTE-SENEGAL-2017-2021-.x23411.pdf

39. World Health Organization (WHO). Global Health Expenditure Database [Internet]. [cited 2024 Aug 10]. Available from: https://apps.who.int/nha/database

40. Organisation for Economic Co-operation and Development (OECD). Creditor Reporting System (online database) [Internet]. [cited 2024 Aug 10]. Available from: https://www.oecd-ilibrary.org/development/data/creditor-reporting-system_dev-cred-data-en

41. Sharma L, Heung S, Twea P, Yoon I, Nyondo J, Laviwa D, et al. Donor coordination to support universal health coverage in Malawi. Health Policy Plan. 2024;39(Suppl 1):i118.

42. Tadesse AW, Gurmu KK, Kebede ST, Habtemariam MK. Analyzing efforts to synergize the global health agenda of universal health coverage, health security and health promotion: a case-study from Ethiopia. Global Health. 2021;17(1):53.

43. World Bank Group Development Finance. A Changing Landscape: Trends in official financial flows and the aid architecture [Internet]. 2021 Nov [cited 2024 Aug 10]. Available from: https://thedocs.worldbank.org/en/doc/9eb18daf0e574a0f106a6c74d7a1439e-0060012021/original/A-Changing-Landscape-Trends-inOfficial-Financial-Flows-and-the-Aid-Architecture-November-2021.pdf

44. Kohler JC, Bowra A. Exploring anti-corruption, transparency, and accountability in the world Health organization, the United nations development programme, the world bank group, and the global fund to fight AIDS, tuberculosis and malaria. Global Health. 2020;16:1–10.

45. Gorodensky A, Bowra A, Saeed G, Kohler J. Anti-corruption in global health systems: using key informant interviews to explore anti-corruption, accountability and transparency in international health organisations. BMJ Open. 2022;12(12):e064137.

46. García PJ. Corruption in global health: the open secret. The Lancet. 2019;394(10214):2119– 24.

47. Mackey TK, Kohler JC, Savedoff WD, Vogl F, Lewis M, Sale J, et al. The disease of corruption: views on how to fight corruption to advance 21 st century global health goals. BMC Med. 2016;14:1–16.

48. Chang Z, Rusu V, Kohler JC. The Global Fund: why anti-corruption, transparency and accountability matter. Global Health. 2021;17:1–11.

49. McCoy D, Jensen N, Kranzer K, Ferrand RA, Korenromp EL. Methodological and policy limitations of quantifying the saving of lives: a case study of the global fund’s approach. PLoS Med. 2013;10(10):e1001522.

50. Friebel R, Silverman R, Glassman A, Chalkidou K. On results reporting and evidentiary standards: spotlight on the Global Fund. The Lancet. 2019;393(10184):2006–8.

51. Hanefeld J. How have global health initiatives impacted on health equity? Promot Educ. 2008;15(1):19–23.

52. Storeng KT. The GAVI Alliance and the ‘Gates approach’to health system strengthening. Glob Public Health. 2014;9(8):865–79.

53. Biesma RG, Brugha R, Harmer A, Walsh A, Spicer N, Walt G. The effects of global health initiatives on country health systems: a review of the evidence from HIV/AIDS control. Health Policy Plan. 2009 Jul 1;24(4):239–52.

54. Bertone MP, Palmer N, Kruja K, Witter S, 1 HWG. How do we design and evaluate health system strengthening? Collaborative development of a set of health system process goals. Int J Health Plann Manage. 2023;38(2):279–88.

55. The Global Fund. Managing investments in Resilient and Sustainable Systems for Health [Internet]. Geneva; 2019 May [cited 2024 Sep 29]. Available from: https://www.theglobalfund.org/media/8441/oig_gf-oig-19-011_report_en.pdf#:~:text=The%20Office%20of%20the%20Inspector%20General%20(OIG)%20safeguards%20the%20assets

56. The Global Fund Technical Evaluation Reference Group. Global Fund Mapping Health Systems Strengthening (HSS) Component of the Resilient and Sustainable Systems for Health (RSSH) Investments: TERG Position Paper, Management Response, and Final Report [Internet]. 2023 Jun [cited 2024 Aug 10]. Available from: https://www.theglobalfund.org/media/13115/terg_mapping-hss-component-rssh_report_en.pdf

57. Global Oxygen Alliance (website) [Internet]. [cited 2024 Aug 10]. Available from: https://globaloxygenalliance.org/

58. Thornton J. Hepatitis Fund aims to accelerate viral hepatitis elimination. The Lancet. 2023;401(10386):1414–5.

59. World Health Organization (WHO). Health4Life Fund. A global financing partnership on non-communicable diseases and mental health [Internet]. 2022 [cited 2024 Aug 10]. Available from: https://knowledge-action-portal.com/en/news_and_events/news/6559

60. Boyce MR, Sorrell EM, Standley CJ. An early analysis of the World Bank’s Pandemic Fund: a new fund for pandemic prevention, preparedness and response. BMJ Glob Health. 2023 Jan;8(1):e011172.

61. Samarasekera U. New US $1· 5 billion fund for primary care. The Lancet. 2023;402(10397):174.

62. Sachs JD, Binagwaho A, Birdsall N, Broekmans J, Chowdhury M, Garau P, et al. Investing in Development A Practical Plan to Achieve the Millennium Development Goals: Overview. Routledge; 2019.

63. Labonte R, Marriott A. IHP+: little progress in accountability or just little progress? The Lancet. 2010;375(9725):1505–7.

64. Shorten T, Taylor M, Spicer N, Mounier-Jack S, McCoy D. The International Health Partnership Plus: rhetoric or real change? Results of a self-reported survey in the context of the 4 th high level forum on aid effectiveness in Busan. Global Health. 2012;8:1–13.

65. Witter S, Baker P. Tracking Delivery on the Lusaka Agenda. Policy Paper 336 [Internet]. Center for Global Development. Washington, DC; 2024 Sep [cited 2024 Sep 28]. Available from: https://www.cgdev.org/publication/tracking-delivery-lusaka-agenda

66. Drake T, Khan M. Center for Global Development. 2024 [cited 2024 Sep 28]. A New Compact for Health Financing: The Global Political Economy of Reform. Available from: https://www.cgdev.org/blog/new-compact-health-financing-global-political-economy-reform

67. The Global Fund to Fight AIDS Tuberculosis and Malaria. Annual Financial Report 2023 [Internet]. Geneva; 2024 Apr [cited 2024 Aug 10]. Available from: https://www.theglobalfund.org/media/14110/corporate_2023annualfinancial_report_en.pdf

68. Gavi the vaccine alliance. webpage. 2024 [cited 2024 Aug 10]. Funding: Current period 2021– 2025. Available from: https://www.gavi.org/investing-gavi/funding/current-period-2021-2025

69. The Global Financing Facility (GFF). The Global Financing Facility in Support of Every Woman Every Child: Executive Summary [Internet]. [cited 2024 Aug 10]. Available from: https://www.worldbank.org/content/dam/Worldbank/document/HDN/Health/GFF-Executive-Summary_EN.pdf

70. Unitaid. Proposed Operating Expenses Budget 2023 [Internet]. Geneva; 2022 Dec [cited 2024 Aug 10]. Available from: https://unitaid.org/assets/UNITAID_EB41_2022_7_2023-Budget.pdf

71. Unitaid. Unitaid’s Investment Case 2023-2027 [Internet]. Geneva; 2023 [cited 2024 Aug 10]. Available from: https://unitaid.org/assets/Unitaid_InvestmentCase_Report.pdf

72. FIND. FIND Strategy 2021: Testing for healthy and safe lives [Internet]. Geneva; 2021 [cited 2024 Aug 10]. Available from: https://www.finddx.org/about-us/strategy/

73. CEPI. Funding and Expenditure (December 2018) [Internet]. London, UK; 2024 [cited 2024 Aug 10]. Available from: https://static.cepi.net/downloads/2023-12/050319-Funding-and-Expenditure-Final_V3.pdf

74. Pakistan Bureau of Statistics (PBS) G of P. National Health Accounts Pakistan 2019-20 [Internet]. Islamabad; 2022 [cited 2024 Aug 25]. Available from: https://www.pbs.gov.pk/publication/national-health-accounts-pakistan-2019-20

75. WHO Regional Office for the Eastern Mediterranean. Country Private Health Sector Effective Engagement for Service Provision Pakistan. Cairo; 2019.

76. The Global Fund Office of the Inspector General. Audit Report: Global Fund Grants in the Islamic Republic of Pakistan [Internet]. Geneva; 2020 Apr [cited 2024 Aug 25]. Available from: https://www.theglobalfund.org/media/9595/oig_gf-oig-20-012_report_en.pdf

77. Global Financing Facility. Pakistan country profile: Global Financing Facility Data Portal.

78. Direction de la Santé de la Mère et de l’Enfant. Ministère de la Santé et de l’Action Sociale (République du Senégal). Dossier d’investissement pour la réduction de la mortalité maternelle, néo-natale, infanto-juvénile, des adolescents et des jeunes [Internet]. Dakar; 2019 Jun [cited 2024 Sep 29]. Available from: https://www.globalfinancingfacility.org/sites/gff_new/files/documents/Senegal-dossier-dinvestisement.pdf

